# Meta-SAIGE: Scalable and Accurate Meta-Analysis for Rare Variants

**DOI:** 10.1101/2024.09.17.24313855

**Authors:** Eunjae Park, Kisung Nam, Seokho Jeong, Karl Keat, Dokyoon Kim, Vikas Bansal, Wei Zhou, Seunggeun Lee

## Abstract

A meta-analysis is a practical approach to increasing the power of rare variant tests by combining summary statistics from multiple cohorts. However, existing methods for rare variant meta-analysis often fail to correctly control type I error rates when analyzing low-prevalence binary traits and are computationally intensive when analyzing many phenotypes. This paper introduces Meta-SAIGE, a novel approach for rare variant meta-analyses that addresses these challenges. Meta-SAIGE reduces type I error inflation through precise estimation of the distribution of test statistics and allows the reuse of the linkage disequilibrium (LD) matrix across phenotypes, significantly improving computational efficiency for phenome-wide analyses. Simulation studies using UK Biobank whole-exome sequencing (WES) genotypes demonstrate that Meta-SAIGE effectively controls type I error rates and yields power similar to that of pooled individual-level data through SAIGE-GENE+. A meta-analysis of UK Biobank and All of Us WES data for 83 low prevalence disease phenotypes identified 237 associations. Notably, 80 of these associations were not significant in either dataset alone, underscoring the power of our meta-analysis.

## Introduction

The advent of mega biobanks and consortia has significantly enhanced our ability to sequence large cohorts, facilitating the identification of rare genetic variants linked to human diseases and traits. Due to low minor allele frequencies, single-variant tests are underpowered for identifying rare variant associations [1–3]. In response, gene or region-based association tests, which aggregate the effects of multiple rare variants, have been proposed. Different statistical tests such as the Burden test [4], SKAT [5], and SKAT-O [6] have been pivotal in the analysis of rare variant associations. More recently, STAAR [7], SAIGE-GENE [8] and SAIGE-GENE+ [9] have been introduced, offering efficient implementation of rare variant tests for large-scale biobank datasets, while accommodating sample relatedness and case–control imbalances.

Meta-analysis is a practical approach for identifying genetic associations that may not be detectable in individual cohorts but become statistically significant when summary statistics from multiple studies are combined [10]. Meta-analysis is particularly relevant for rare variant association tests, where the low frequencies of the variants can limit test power in individual studies. Substantial efforts have been made to facilitate the meta-analysis of rare variants association studies. For example, international consortia such as the Biobank Rare Variant Analysis (BRaVa) have been established. As more biobanks carry out sequencing studies, rare-variant meta-analysis is likely to become a primary approach for identifying novel rare variant associations for human diseases and traits.

Several methodologies for rare variant meta-analyses have been developed, notably RareMetal [11, 12] and MetaSKAT [1]. More recently, MetaSTAAR [13] was introduced, enhancing the analysis by integrating variant functional annotations and accommodating sample relatedness. However, MetaSTAAR can exhibit significantly inflated type I error rates under imbalanced case–control ratios (Supplementary Note 1 and Supplementary Figure 1), a prevalent challenge in biobank-based disease phenotype studies. Additionally, MetaSTAAR’s requirement for constructing separate linkage disequilibrium (LD) matrices for each phenotype substantially increases the computational load when analyzing multiple phenotypes. An alternative approach is the Fisher method [14], which combines gene or region test p-values from each cohort, but it often has substantially lower power than joint analysis with individual-level data.

This paper proposes a new rare variant meta-analysis method, Meta-SAIGE, which extends SAIGE-GENE+ [9] to meta-analysis. First, Burden, SKAT, and SKAT-O test p values are calculated using per-variant score statistics and LD matrices from each cohort. Next, to address type I error inflation in the presence of case–control imbalance, Meta-SAIGE employs two-level saddlepoint approximation (SPA) [15, 16], including SPA on score statistics of each cohort, and a genotype count-based SPA for combined score statistics from multiple cohorts [17]. In addition, Meta-SAIGE allows the use of a single sparse LD matrix across all phenotypes, significantly reducing computational costs when conducting phenome-wide analyses involving hundreds or thousands of phenotypes. Simulation studies using UK Biobank (UKB) whole-exome sequencing (WES) data [18] demonstrate that Meta-SAIGE effectively controls type I error rates and is computationally faster and more efficient than MetaSTAAR. We conducted a meta-analysis of UKB and All of Us [19] exome-wide rare-variant gene-based tests for data for 83 disease phenotypes and identified 237 associations at the exome-wide significance level. Among these associations, 80 were not significant in either dataset alone, underscoring the power of our meta-analysis.

## Results

### Overview of Meta-SAIGE

Meta-SAIGE contains three steps: (1) preparing per-variant level association summaries and a sparse LD matrix for each cohort, (2) combining summary statistics from all the studies into a single superset, and (3) running gene-based tests, as illustrated in Figure 1.

**Fig. 1.**
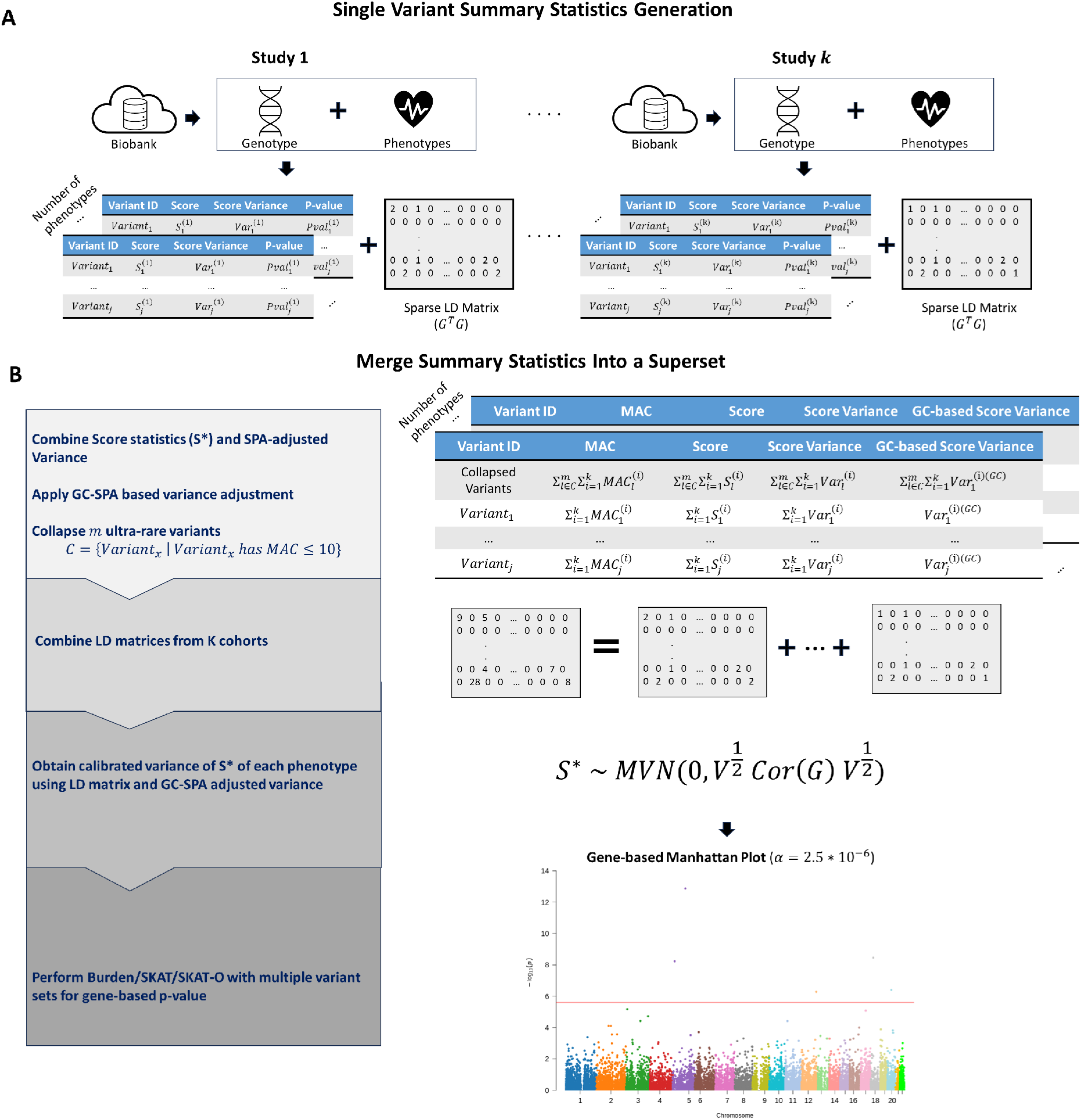
Overview of Meta-SAIGE. (A) Summary statistics generated by each cohort for Meta-SAIGE. (B) Meta-analysis incorporating per-variant summary statistics (score statistics *S* and its variance *V*) and sparse LD matrices. The variance *V* is adjusted using SPA. Multiple summary statistics are merged into a superset, with additional variance adjustments via the GC-based SPA method. Ultrarare variants are collapsed, followed by gene-based tests using various functional annotations and minor allele frequency (MAF) cutoffs. P-values from these tests are combined using the Cauchy combination method.

The first step uses SAIGE [16] to derive per-variant score statistics, *S*, accompanied by their variance and association p-values. SAIGE conducts the score test based on a generalized linear mixed model that can produce accurate single variant p-values by adjusting for case–control imbalance and sample relatedness (Figure 1. **A**). For binary phenotypes, the computation of p-values involves two possible methods, saddlepoint approximation (SPA) and efficient resampling [20], depending on the minor allele count (MAC). Additionally, the sparse LD matrix, Ω, is generated in each cohort, which is the pairwise cross product of dosages across genetic variants. In contrast to MetaSTAAR, which uses a phenotype variance weighted LD matrix, the sparse LD matrix used by Meta-SAIGE is not phenotype specific, allowing us to reuse the same LD matrix across different phenotypes. When meta-analyzing *M* variants from *K* cohorts, the storage requirement for a single phenotype for the sparse LD matrix is *O*(*MFK*), where *F* is the number of variants with the nonzero cross-product on average, and for single variant summary statistics, it is *O*(*MK*). When the meta-analyses are performed for *P* different phenotypes, our approach requires *O*(*MFK* + *MKP*) storage, while MetaSTAAR requires *O*(*MFKP* + *MKP*) storage. Next, score statistics from multiple cohorts are consolidated into a single statistic (Figure 1. **B**). For binary traits, the variance of each score statistic is recalculated by inverting the p-value by SAIGE-GENE+, which is a method that we previously developed for rare-variant association test. To further improve type I error control, we applied the Genotype-Count (GC) based saddlepoint approximation [17], which was previously designed to improve type I error control in the meta-analysis setup. The covariance matrix of score statistics is calculated in a sandwich form, *Cov*(*S*) = *V* ^1*/*2^*Cor*(*G*)*V* ^1*/*2^. Here *Cor*(*G*) is the correlation matrix of genotypes and is calculated with the sparse LD matrix Ω, and *V* is the diagonal matrix of the variance of *S*, which is obtained by inverting the SPA-GC-adjsuted p-values. With the combined per-variant statistics and covariance matrix, Meta-SAIGE performs rare-variant tests in the same manner as SAIGE-GENE+. It conducts Burden, SKAT, and SKAT-O set-based tests utilizing various functional annotations and MAF cutoffs. The ultrarare variants (e.g. MAC *<* 10) are collapsed to enhance type I error control and power, while reducing computation cost as demonstrated in SAIGE-GENE+. The Cauchy combination method [21, 22] is then used to combine p-values corresponding to different functional annotations and MAF cutoffs for each testing gene or region.

### Type 1 Error and Power Evaluation

To evaluate the type I error rates of Meta-SAIGE, simulations were conducted using the UKB WES data of 160,000 white British participants, which were partitioned into three non-overlapping cohorts of varying sizes. Null phenotypes were generated for each cohort, with prevalence of 5% and 1%. The simulation process was replicated 60 times to yield approximately one million tests.

The empirical type I error rates are presented in Figure 2 and Supplementary Table 1. Without any adjustment, mirroring MetaSTAAR, there was a significant inflation in type I error rates. For example, with a disease prevalence of 1% and a sample size ratio of 1:1:1, the type I error rate for the no-adjustment method at *α* = 2.5*×*10^*−*6^ was *α* = 2.12*×*10^*−*4^, nearly 100 times higher than the nominal level (Supplementary Table 1). The SPA applied to score statistics of each cohort (SPA adjustment) notably decreased type I error rates, albeit with some remaining inflation. Meta-SAIGE, which further applies a GC-based SPA, outperformed other methods, demonstrating relatively well controlled type I error rates. Based on the same UKB genotype data, we further conducted simulation to assess the power of Meta-SAIGE in detecting rare-variant associations (Figure 3 and Supplementary Table 2). We examined various scenarios of different effect sizes of rare variants and benchmarked Meta-SAIGE’s performance against a joint analysis of individual-level data via SAIGE-GENE+ and the weighted Fisher’s method [14], which aggregates SAIGE-GENE+ p-values weighted by sample size. Meta-SAIGE consistently demonstrated statistical power on par with that of joint analyses across all evaluated scenarios. In contrast, the weighted Fisher’s method yielded significantly lower power, underscoring the potential of our approach to enhance the detection of rare variant associations.

**Fig. 2.**
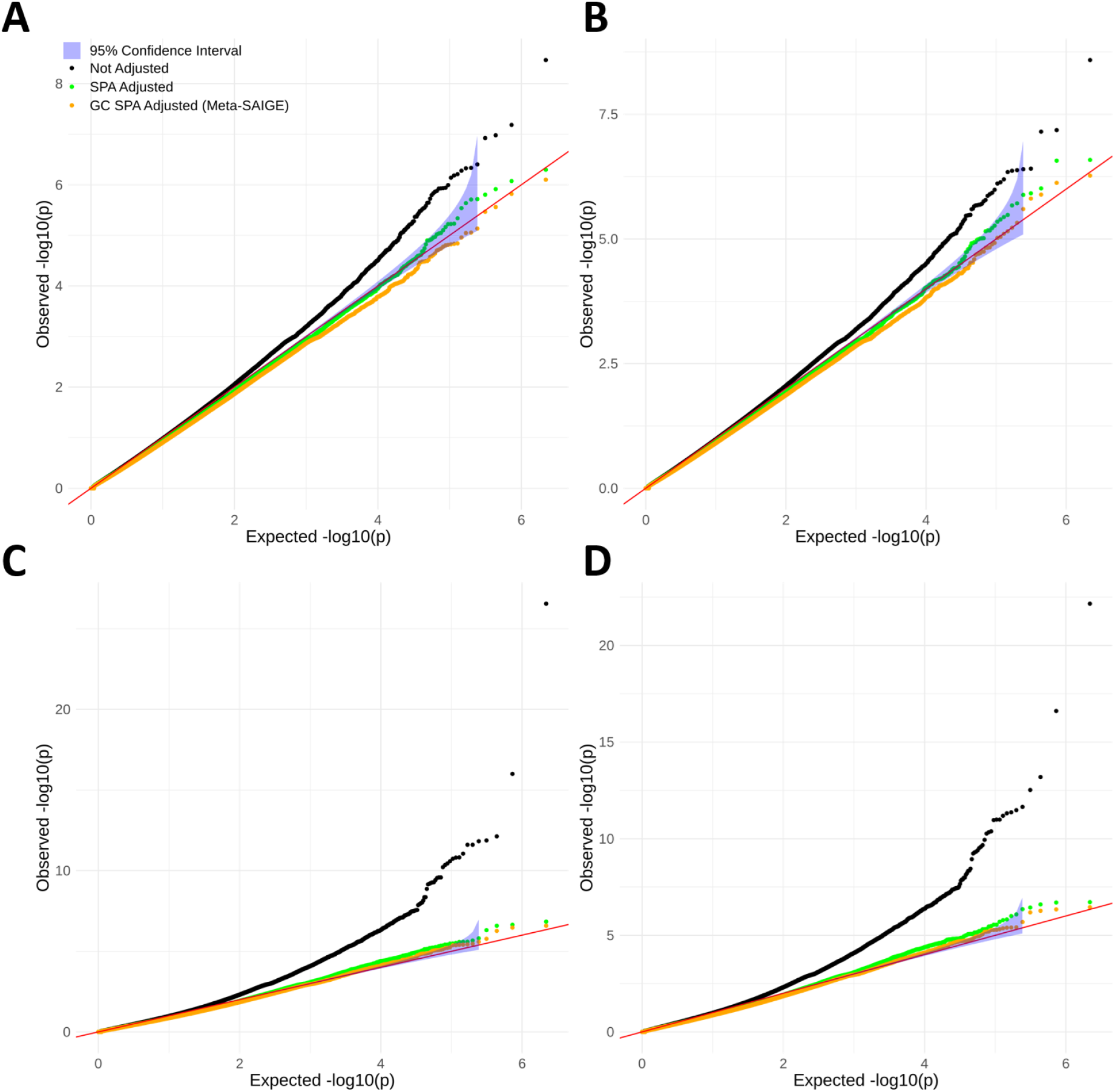
QQ-plots for null phenotypes with prevalence of 5% and 1% with different sample size ratios (1:1:1 and 4:3:2). (A) QQ-plot for a 1:1:1 sample size ratio with prevalence=5%. (B) QQ-plot for a 4:3:2 sample size ratio with prevalence=5%. (C) QQ-plot for a 1:1:1 sample size ratio with prevalence=1%. (D) QQ-plot for a 4:3:2 sample size ratio with prevalence=1%.

**Fig. 3.**
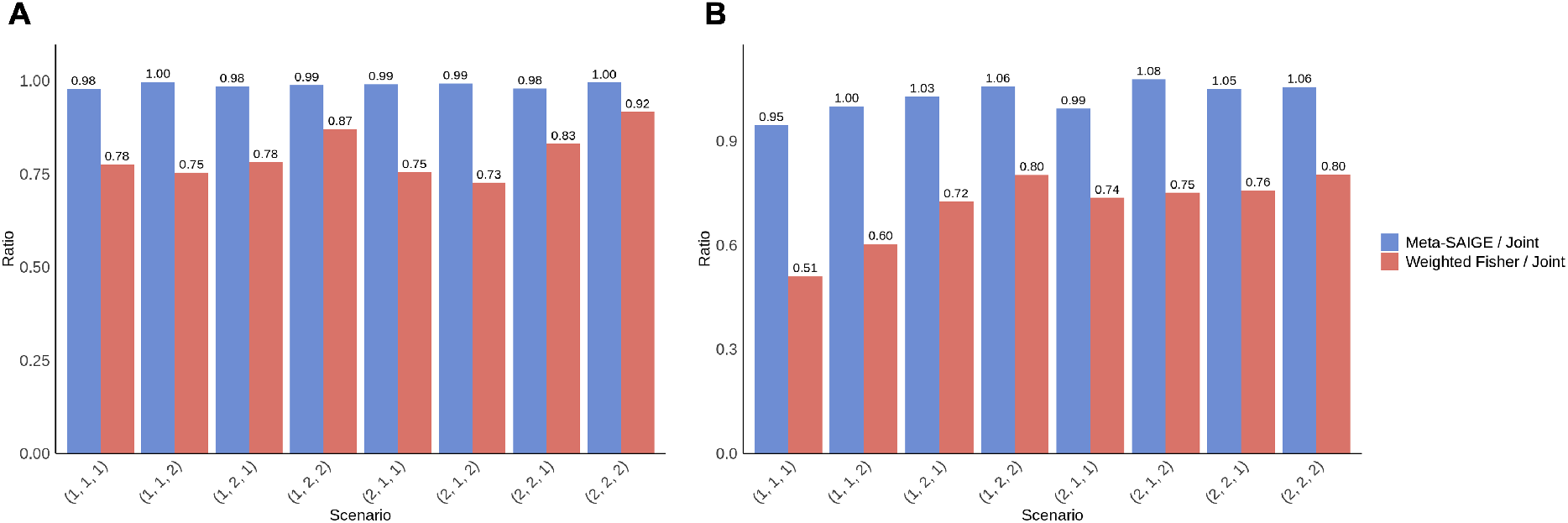
Power evaluation results for the binary phenotype with (A) 5% prevalence and (B) 1% prevalence. The scenarios are annotated with three indices, (*i, j, k*), which indicate the proportion of causal variant (*i*), magnitude of the effect sizes (*j*), direction of the effect of causal variants (*j*). Details can be found in Supplementary Table 2. The Y-axis represents the power ratio, with a value of one indicating equal power compared to the joint test.

### Computational Cost Evaluation

We conducted a comparative analysis of the computational costs of Meta-SAIGE and MetaSTAAR. Because MetaSTAAR software lacks the ability to read genotype data directly, we utilized seqminer[23] for genotype data preprocessing. Figure 4 shows the computation time and memory usage for analyzing the largest gene, *TTN* (number of rare variants = 17,577, number of ultrarare variants = 15,098), across three key meta-analysis tasks: per-variant statistics generation, LD matrix generation, and the meta-analysis itself. The first two tasks should be performed independently for each cohort.

**Fig. 4.**
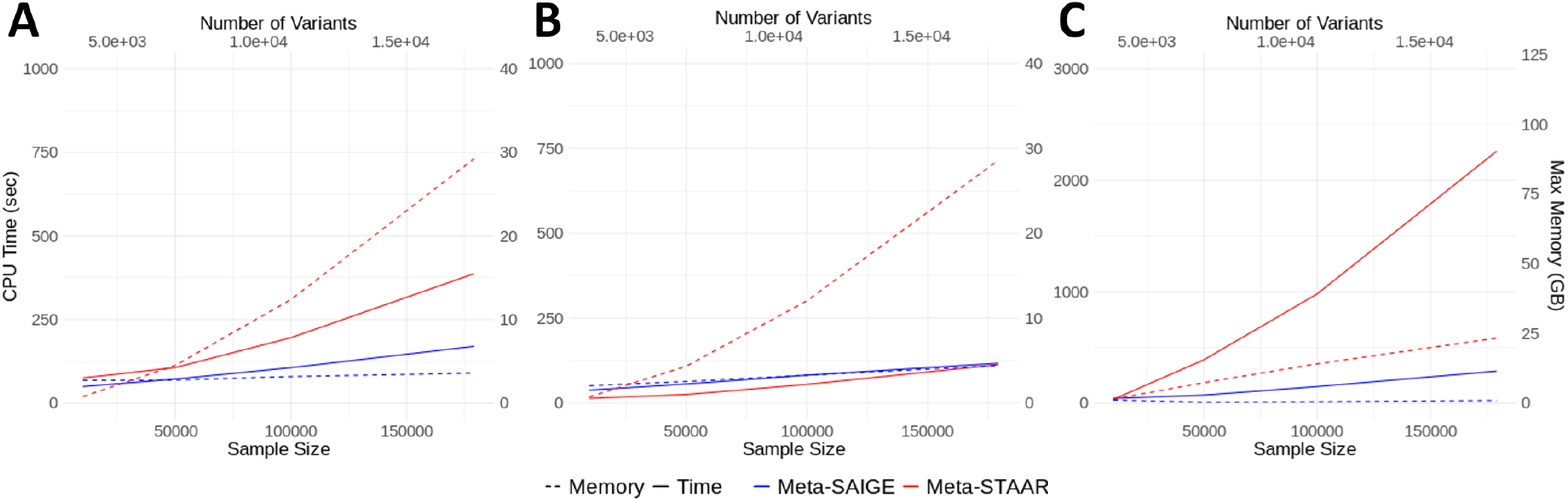
Computational cost evaluation results on *TTN*. (A) Computation time for input file generation, including null model fitting and summary statistics generation. (B) Computation time for LD matrix generation. (C) Computation time for the meta-analysis step.

Our results indicate that Meta-SAIGE consistently results in lower computational demands in terms of both CPU time and memory usage. For example, with a sample size of 179, 176, Meta-SAIGE was significantly faster than MetaSTAAR, with speed improvements of 8.1, 6.5, and 24.7 times for per-variant statistics generation, LD matrix generation, and meta-analysis, respectively. The difference in computational efficiency between Meta-SAIGE and MetaSTAAR diminished when smaller genes, such as *MSH6* (number of rare variants = 990), were analyzed (Supplementary Figure 2).

As previously mentioned, MetaSTAAR requires phenotype-specific LD information. Consequently, when the *P* phenotypes were analyzed, the LD matrix (Figure 4. B) must be generated *P* times for each cohort. The storage sizes of the LD matrix for whole-exome sequencing (WES) using Meta-SAIGE were 3.0 GB for UKB and 2.8 GB for All of Us. However, the projected LD matrix storage values for MetaSTAAR were 8.47 GB for UKB and 7.91 GB for All of Us. When 100 phenotypes were analyzed, the storage cost of LD matrices required by MetaSTAAR is projected to be 847 GB for UKB and 791 GB for All of Us, which is about 282 times higher than Meta-SAIGE. Therefore, by using single LD matrix for all phenotypes,Meta-SAIGE significantly reduces the storage costs.

### Meta-analysis of UKB and All of Us Datasets

To illustrate the performance of Meta-SAIGE in real data, we applied Meta-SAIGE to conduct a whole-exome meta-analysis for 83 disease phenotypes using UKB and All of Us European Ancestry samples. Our analysis included 459K European Ancestry samples from UKB and 116K European Ancestry samples from All of Us. Traits were selected to be compatible with both UKB and All of Us with sufficient case sizes while incorporating traits from previous WES phenome-wide analyses[9]. Case–control ratios varied from 1:2 (essential hypertension, PheCODE=401.1) to 1:305 (hemorrhage from gastrointestinal ulcers, PheCODE=531.1) across all analyzed phenotypes. A detailed list of phenotypes is provided in Supplementary Table 3. We applied three MAF cutoffs (1%, 0.1%, 0.01%) and three functional annotations (LoF, LoF+Missense, LoF+Missense+Synonymous), resulting in a total of nine different test masks.

Figure 5 A and Supplementary Table 4 present all significant associations at the exome-wide significance level of *α* = 2.5*×*10^*−*6^ among, with 20,000 protein coding genes tested. Across all the 83 phenotypes, 237 gene–phenotype pairs were identified as significant. The estimated false discovery rate (FDR) was 0.0175 (FDR = 83 [number of traits] *×* 20,000 [number of genes] *×*2.5 *×* 10^*−*6^ [exome-wide significance threshold]/237 [number of significant genes]). When a more stringent criterion adjusted for the number of phenotypes at the top of the exome-wide significance level (p-value *<* 2.5 *×* 10^*−*6^*/*83 = 3.0 *×* 10^*−*8^) was applied, 150 gene–phenotype associations remained significant.

**Fig. 5.**
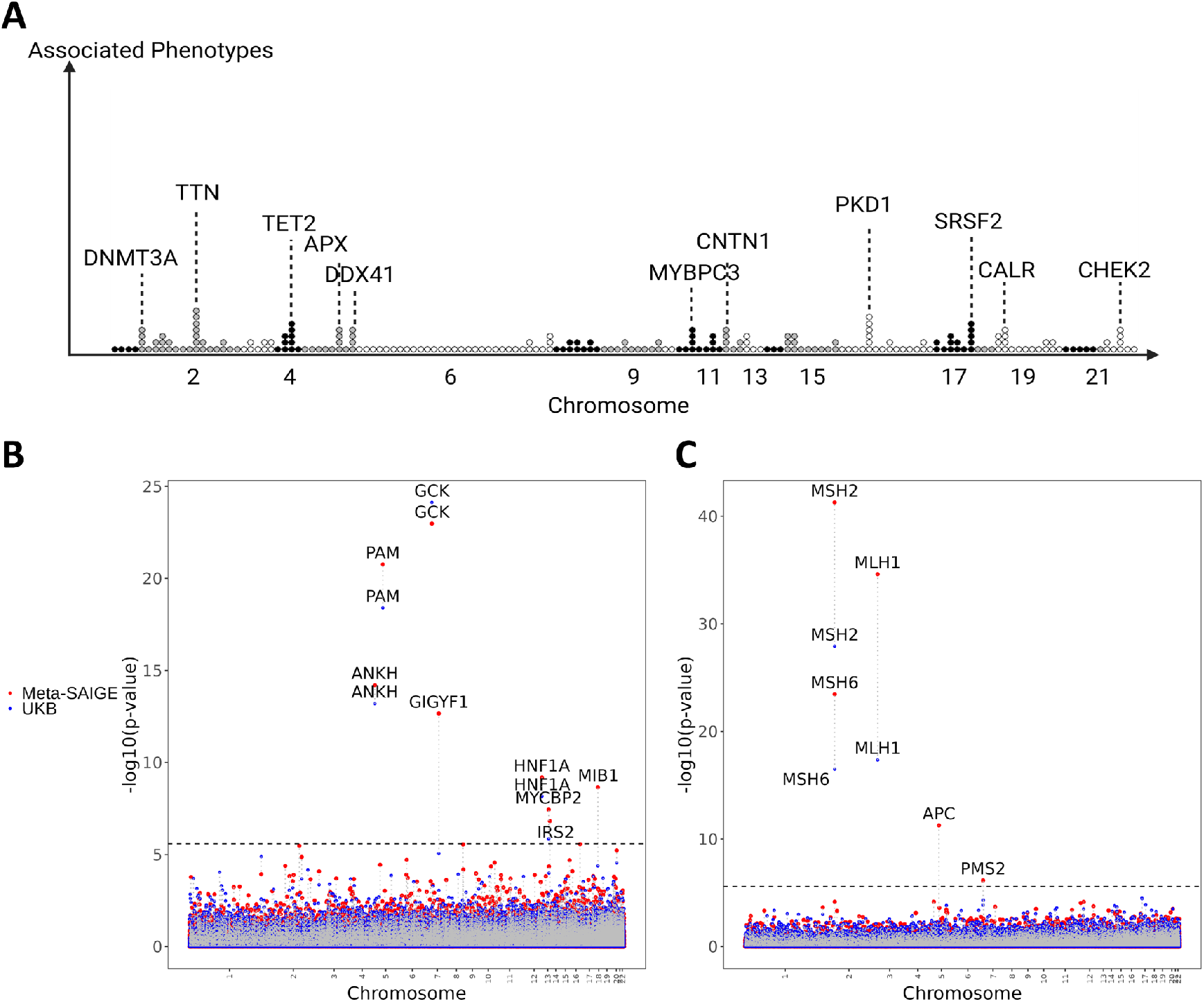
UKB + AllofUs meta-analysis results. (A) Manhattan plot displaying the number of significantly associated traits. (B) Manhattan plot for type 2 diabetes (PheCode 250.2). (C) Manhattan plot for colorectal cancer (PheCode 153). In (B) and (C), p-values calculated using UK Biobank (UKB) data alone are shown in blue, while p-values from Meta-SAIGE combining UKB and All of Us data are shown in red.

Among the 237 associations, 139 and 46 of them were significant in the UKB data and All of US data only, respectively, underscoring the power of the meta-analysis. Among all significant genes, *TTN* and *PKD1* were the most pleiotropic one, being associated with 7 phenotypes, followed by *CHEK2, TET2*, and *SRSF2*, each associated with 5 phenotypes (Figure 5. **A**).

To illustrate the type I error control and improvement in p-values through meta-analysis, we generated Manhattan plots for two phenotypes: type 2 diabetes (T2D) and colorectal cancer (Figure 5 B). A total of five genes were significant for colorectal cancer, and 8 genes were significant for T2D. Our results clearly demonstrate that the meta-analysis can substantially improve the power to detect disease associated genes. For example, Adenomatous polyposis coli (*APC*) is a tumor suppressor gene and defects in this gene increase the risk of colorectal cancer [24]. In our analysis, the association of *APC* with colorectal cancer was not significant in UKB or All of US data alone (UKB p-value=4.40 *×* 10^*−*3^ and All of US p-value=1.32 *×* 10^*−*5^), and the meta-analysis p-value was 5.39 *×* 10^*−*12^, which was much more significant than in individual biobanks.

## Discussion

This study introduces Meta-SAIGE, a method for meta-analyzing rare variant association studies. By employing SPA and GC-based adjustment methods, Meta-SAIGE accurately calculates p-values, effectively controlling type I error rates while preserving true signals. As demonstrated in the type I error simulation, substantial reductions in type I error rates were observed compared to the unadjusted approach. Additionally, Meta-SAIGE improves computation efficiency by allowing the use of a single LD matrix for different phenotypes, facilitating more cost-effective phenome-wide analyses as demonstrated through a meta-analysis of UKB and All of Us WES data.

Our simulation results showed that the meta-analysis based on Meta-SAIGE cohort-level summary statistics by Meta-SAIGE has similar study power to the joint analysis using SAIGE-GENE+ on individual-level data. In real data analysis of UKB and All of Us WES data, highly concordant p-values were observed between the two approaches for three example phenotypes (Supplementary Note B) and both approaches revealed identical significant genes.

Many biobanks are now collecting samples from diverse ancestries to mitigate the Eurocentric bias prevalent in genetic association studies, emphasizing the need for multi-ancestry meta-analysis. Although our study focused on a single ancestry, Meta-SAIGE can also be applied to multi-ancestry samples. A key challenge in multi-ancestry analysis is collapsing ultra-rare variants, as these variants can differ significantly between ancestry groups. To address this, we added a step for ancestry-specific collapsing (see Methods section). For T2D and colorectal cancer, we conducted a multi-ancestry meta-analysis of all European ancestry samples (EUR, n=572K) from UKB and All of Us, as well as African ancestry (n=46K) and American (n=36K) samples from All of Us. Only variants with a minor allele frequency (MAF) *≤* 0.01 across all ancestry groups were included for meta-analysis (Supplementary Figure 4). Supplementary Table 6 lists the significant genes identified by either European-only Meta-analysis (EUR-Meta) or Meta-analysis of Cross Ancestry (Meta-CrossAncestry). Meta-CrossAncestry had generally lower p-values than EUR-Meta and identified two additional associations for colorectal cancer: RNU1-31P and RPL29P11. RPL29P11, Ribosomal Protein L29 Pseudogene 11, is pseudogene of the ribosomal protein L29, which is shown to up-regulate colorectal cancer [25, 26], demonstrating the enhanced power of multi-ancestry meta-analysis. However, a more comprehensive evaluation of its performance across broader and more varied ancestry groups is required, which we leave for future investigation.

Overall, we demonstrated that Meta-SAIGE can effectively handle large-scale rare-variant metaanalyses of binary phenotypes, adjusting for phenotype imbalance. As many biobanks are currently generating large-scale sequencing data, our method can facilitate more powerful meta-analysis studies for identifying rare disease-associated genetic variants and elucidating the genetic architecture of underlying complex traits.

## Methods

### Workflow of Meta-SAIGE

Meta-SAIGE comprises three steps: (1) preparation of single-variant association summaries for each cohort, (2) consolidation of the summaries from all studies into a singular comprehensive dataset, and (3) implementation of gene-based association testing, as illustrated in Figure 1.

### Summary Statistics from Each Cohort

The first step can be performed using SAIGE software to compute per-variant score statistics (*S*), variance (*V*), p-value, and a sparse LD-matrix (Ω). For binary phenotypes, case–control imbalance is accounted for using the SPA test applied to a logistic regression model as detailed by Dey et al.[27].

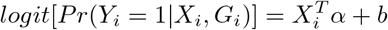

Here, *Y*_*i*_ represents the phenotype of the *i*^*th*^ sample, *X*_*i*_ represents the nongenetic covariates and *b* represents a random effect in the logistic mixed model. Note that for continuous traits, alinear mixed model is used instead. With *K* cohorts, the score statistic for each variant *j* in the *k*^*th*^ cohort is computed as

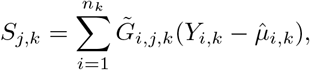

where *n*_*k*_ is the sample size of the *k*^*th*^ cohort, 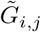 is the covariate adjusted genotype, and 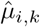 is the estimate of *P r*(*Y*_*i*_ = 1|*x*_*i*_) in the *k*^*th*^ cohort under *H*_0_. The variance of *s*_*j,k*_ (i.e. *v*_*j,k*_) is calculated as

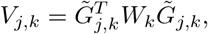

where *W*_*k*_ is a diagonal matrix with 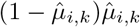 being the *i*^*th*^ diagonal element.

Because phenotype imbalance can lead to an inflation of type 1 error, saddlepoint approximation (SPA) [15] has been proposed to calculate accurate p values. The SPA uses a cumulant generating function (CGF), which can be defined as follows:

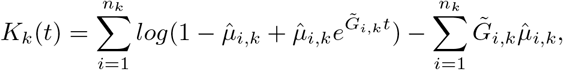

and its second derivative can be used to calculate the p value of each variant. Now, the SPA-adjusted variance, 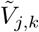, is calculated by inverting the SPA p value with the given score statistic *S*_*j,k*_. The sparse LD matrix (Ω) can also be generated as 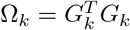

### Combining Summary Statistics Into a Single Super-Set

In Meta-SAIGE, summary statistics from multiple studies are consolidated into a single table, namely, a superset. The superset is subsequently used to perform gene-based tests. In this step, *S*_*j*_ and *V*_*j*_ from each study are combined as

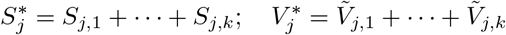

Suppose that a region with the first *m* variants (*j* = 1, …, *m*) is tested. To obtain the covariance matrix of 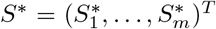, we first calculate the correlation matrix of *S, Cor*, using the sparse LD matrix Ω_*k*_ and the MAF of each variant obtained from each respective cohort.

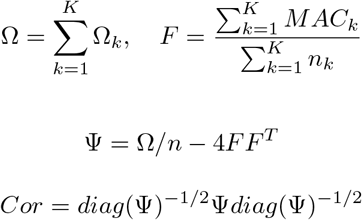

Here *MAC*_*k*_ represents the vector of minor allele counts in cohort k. Note that Ψ is the covariance matrix of *S*^*\**^ and *diag*(Ψ) is a diagonal matrix of Ψ. Then, *S*^*\**^ follows a multivariate normal distribution, given that *Cor* is the correlation among genetic markers

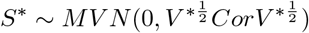

where *V* ^*\**^ is a diagonal matrix with the diagonal element being 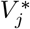. We note that the unadjusted approach uses the unadjusted variance *V*_*j*,1_ to calculate 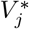

### Gene-Based Test with the Super-Set

Prior to running the region-based test, ultrarare variants are collapsed. Ultrarare variants were determined using the overall MAC calculated from all cohorts, specifically when the overall MAC *≤* 10. The collapsing process aligns with the principles of the burden test, aggregating summary statistics 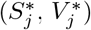 for the selected variants.

Subsequently, gene-based tests are performed. The test statistics *Q* for Burden and SKAT are as follows:

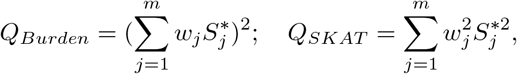

where *w*_*j*_ represents an MAF-based weighting vector calculated from the beta density function with

*Beta*(1, 25). The SKAT-O test is subsequently performed by combining the *Q*_*Burden*_ and the *Q*_*SKAT*_.

For each gene, multiple tests were conducted with various MAF cutoffs and functional annotations. P-values are combined using the Cauchy combination method[21]. In this study, MAF cutoffs of 1%, 0.1%, and 0.01% and functional annotations of, LoF (loss of function), LoF+Missense, and LoF+Missense+synonymous were used.

### Further Adjustment with a GC-based Meta-Analysis Approach

The SPA-adjustment previously outlined assumes that score statistics approximate a normal distribution following variance adjustment using SPA. However, for significantly imbalanced binary phenotypes, this adjustment alone does not suffice to maintain type I error rates within acceptable bounds. The cause of this issue lies in the discrete nature of study-specific statistics, for which mere aggregation proves inadequate. To address this, we previously introduced a genotype-count (GC)-based procedure that uses a GC-based SPA as a reference distribution instead of a normal distribution for meta-analysis [17].

The GC-based method estimates the cumulant generating function of the aggregated score statistics 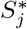 utilizing genotype counts to better approximate 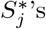 distribution, thereby enhancing the accuracy of the p-value estimation. This procedure is specifically applied to calculate the p value of 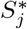 when the single variant p value, derived from variance adjustment, falls below 0.05. We applied it to calculate the p value of 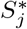 when the single variant p value calculated from the variance adjustment was smaller than 0.05. 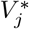 is subsequently determined by inverting the p-value obtained through the genotype-count (GC) based approach. Simulation studies demonstrate that this GC-based meta-analysis approach markedly diminishes type I error by providing a more accurate estimate of the score distribution.

### Ancestry-Specific Collapsing

Given that ultra-rare variants are population-specific, Meta-SAIGE can perform ancestry-specific ultrarare variant collapsing for multi-ancestry meta-analysis. For cohorts sharing the same ancestry, ancestry-specific collapsing is conducted prior to creating the super-set. These ancestry-specifically collapsed variants are treated as unique pseudo-variants.

### Type 1 Error and Power Evaluation

For both type I error and power simulations, we used WES genotype data from 166,960 white British samples in UKB. For type I error evaluation, null phenotypes were generated using the same procedure described in [9]. The logistic mixed model was used to generate the null phenotypes,

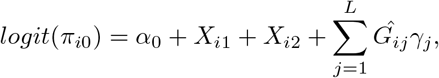

where α_0_ is the intercept term determined by the phenotype prevalence (5% and 1%); *X*_*i*1_ and *X*_*i*2_ are simulated binary and continuous covariates from Bernoulli(0.5) and *N* (0, 1), respectively; 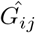 is the standardized genotype value of the ith individual and jth marker, and *γ*_*j*_ is the genetic effect size from *N* (0, 1*/L*). These markers represent polygenic effects. Only even chromosome markers were used for null phenotype generation if the analysis was conducted on odd chromosomes and vice versa. A total of *L* = 30, 000 LD pruned markers were used.

Null phenotype generation was replicated 60 times to yield approximately one million tests. The samples were divided into three cohorts with sample sizes designed to reflect specific ratios. Related samples (pairwise values in GRM *≥* 0.05) were assigned to the same cohort. In the first scenario, samples were distributed relatively equally across three cohorts, assigning (55,655, 55,654, and 55,652) samples, respectively. In the second scenario, a 4:3:2 ratio was used, with values of (74,205, 55,654, 37,102).

To assess the statistical power, we simulated binary phenotypes using ten specific genes. Specifically, a phenotype was generated from the following model,

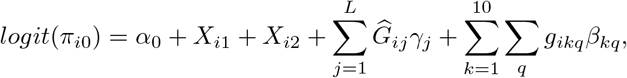

where *π*_*i*0_ is the probability for the *i*th individual being a case given the covariates and genotype, 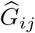 is the standardized genotype value of the *j*th variant of the *i*th individual, *β*_*j*_ is the genetic effect size following *N* (0, 1*/L*), *g*_*ikq*_ is the genotype value of the *q*th rare variant in gene *k* of the *i*th individual, and *β*_*kq*_ is the genetic effect size of the *q*th rare variant in gene *k* according to each scenario described below. Like in the type I error simulation, two covariates *X*_*i*1_ and *X*_*i*2_ were generated from Bernoulli(0.5) and *N* (0, 1), respectively, and the intercept *α*_0_ was determined to set the overall disease prevalences at 1% and 5%.

Our investigation encompassed eight distinct scenarios, each characterized by the proportions of causal variants, effect size magnitudes, and effect directions of rare variants. Specifically, we considered two different configurations of causal variant proportions for non-ultra-rare (MAC *>* 10) variants: (1) 20% of LoF, 10% of missense, and 2% of synonymous and (2) 30% of LoF, 20% of missense and 2% of synonymous. Given that functionally deleterious variants are more likely to be rare, we posited that the proportions of causal variants among ultrarare variants (MAC *≤* 10) were threefold greater than those among less rare variants across various functional annotations. We explored three different settings for absolute effect sizes: (1) |0.5 log_10_(*MAF*)| for LoF and |0.25 log_10_(*MAF*)| for missense and synonymous and (2) |0.7 log_10_(*MAF*)| for LoF and |0.35 log_10_(*MAF*)| for missense and synonymous. We also examined two alternative configurations for effect directions among causal variants: (1) all causal variants exhibited the same directional effect, and (2) 100% LoF, 80% missense, and 50% synonymous variants increased the risk of disease, whereas the remaining causal variants decreased the risk of disease. For each simulation scenario, we conducted 100 independent repetitions, resulting in a total of 1,000 gene-level p-values. We partitioned 166,960 White British samples evenly distributed across three cohorts, assigning 55,655, 55,654, and 55,652 samples, respectively.

### UKB and All of Us WES Data Meta-analysis

We performed a meta-analysis on data from the UKB and All of Us datasets for 83 binary traits among individuals of European ancestry (Supplementary Table 3). Binary traits were phenotyped based on Phecodes [28], which were derived from ICD records. Per-variant summary statistics and LD matrix calculations and meta-analyses were performed on the UKB Research Analysis Platform and the All of Us Workbench cloud system.

WES data from 459,369 white UK Biobank participants were used to create association summaries. Hospital inpatient data coded in the ICD-10 were transformed into Phecodes, and participants were categorized into case–control groups accordingly. PLINK binary files of whole-exome sequences were used for the analysis. Genomic and electronic health record (EHR) data from All of Us were retrieved from the v7 Curated Data Repository (CDR) on the controlled tier. The multiancestry grouping of All of Us participants was derived from the Global Diversity Array (GDA), where principal components were used for ancestry labels [19]. Whole genome sequencing data from All of Us in BGEN format were used to create per-variant summary statistics. EHR data based on ICD-9 and ICD-10 codes were used to map Phecodes. Samples without EHR data were excluded from the analysis.

For both the UKB and All of Us datasets, we included age, sex, batch information, and 10 ancestry principal components as covariates. Summary statistics from the UK Biobank and All of Us were generated with the maximum minor allele frequency of 0.01. To create the linkage disequilibrium (LD) matrix, variants pooled from both cohorts were annotated using VEP105 + LOFTEE GRCh 38 v. 1.04 (https://github.com/BRaVa-genetics/vep105_loftee) into three functional categories: LoF, missense, and synonymous variants.

Additionally, we performed a meta-analysis on multiple ancestry groups across UKB and All of US on Type 2 diabetes and colorectal cancer. Multi-ancestry grouping of All of US participant is derived from Global Diversity Array(GDA), where principal components were generated and projected onto the ancestry label provided by GnomAD. Ancestry groups of over 10,000 samples from All of Us, European(EUR, n = 116,566), African(AFR, n = 45,772), and American(AMR, n=36,152), were used for this analysis. Per-variant summary statistics and LD matrix of UKB-White, All of Us EUR, AFR, and AMR groups were generated using SAIGE and applied to Meta-SAIGE.

## Data Availability

Meta-SAIGE is implemented as an open-source R package available at https://github.com/leelabsg/META_SAIGE. Single variant summary statistics and LD matrix can be generated from SAIGE available at https://github.com/saigegit/SAIGE. UKB and All of US European meta-analysis results are available at https://meta-saige.leelabsg.org/.

## Computational Cost Evaluation

We evaluated the computational cost of MetaSTAAR and Meta-SAIGE across different sample sizes: 10,000, 50,000, 100,000, and 179,176 samples. Each sample size was further divided into three sub-cohorts. For comparison, we primarily used the TTN gene, the largest gene, which represents a computational bottleneck for the analysis. We included LoF, missense, and synonymous variants with a minor allele frequency (MAF) *≤* 0.001, resulting in a total of 17,577 variants. Additionally, we analyzed a smaller gene, MSH6, which had 990 variants. We measured CPU time and memory usage across three distinct stages of the analysis: (1) input file generation, including fitting the null model and conducting single variant association tests; (2) linkage disequilibrium (LD) matrix generation; and (3) meta-analysis.

## URLS

- Meta-SAIGE: https://github.com/leelabsg/META_SAIGE
- SAIGE: https://github.com/saigegit/SAIGE
- UKB+AllofUs meta-analysis PheWeb: https://meta-saige.leelabsg.org/
- SPAtest: https://github.com/leeshawn/SPAtest
- VEP105 + LOFTEE GRCh 38 v. 1.04 : https://github.com/BRaVa-genetics/vep105_loftee

## Inclusions and Ethics Statement

Our study adhered to ethical conduct and inclusive practices throughout. All collaborators meeting Nature Portfolio journals’ authorship criteria are included as authors. We utilized data from All of Us and UKB, respecting their established data access protocols. Our research complied with relevant ethical regulations for secondary data analysis and posed no additional risks to participants. We implemented rigorous data management and analysis protocols. We maintained responsible practices in data interpretation, acknowledging the strengths and limitations of the datasets used.

## Acknowledgements

This research was supported by the Brain Pool Plus (BP+, Brain Pool+) Program through the National Research Foundation of Korea (NRF) funded by the Ministry of Science and ICT (2020H1D3A2A03100666). We thank the helpful discussions from Dr. Duncan Palmer and the “Biobank Rare Variant Analysis” (BRaVa) consortium. UK Biobank data were accessed under the accession number UKB: 45227.

## Author Contributions

E.P. and S.L. implemented the Meta-SAIGE software. E.P. conducted the evaluation of type 1 error inflation. K.N. carried out the power evaluation. S.J. conducted meta-analysis using data from the UK Biobank and All of Us, with input from D.K. and K.K. Helpful advice was provided by D.K. and V.B. The manuscript was written by E.P., K.N., S.J., W.Z., and S.L.

