## Supplementary Materials for "Meta-SAIGE: Scalable and Accurate Meta-Analysis for Rare Variants"

**A. MetaSTAAR Simulation Result**

The MetaSTAAR method was evaluated through a simulation study based on the UKB WES data, focusing exclusively on White British samples. The sample was divided into three cohorts, each comprising 10,000 unrelated individuals. Binary phenotypes were randomly generated with a prevalence of 1%. *FAM83F* gene from chromosome 22, which contains approximately 168 variants, was selected for analysis. This simulation was repeated 100 times. **Supplementary Figure 1** shows the QQ plot of the analysis.

**B. Comparison of Meta-SAIGE to SAIGE-GENE+**

We further evaluated Meta-SAIGE using real binary phenotypes with low prevalences. Type 2 diabetes (T2D), which has a prevalence of 5%, was selected along with glaucoma and colorectal cancer (ColCa), each with a prevalence of 1%. The phenotypes are identified by their respective Phecodes as T2D (250.2), glaucoma (365), and ColCa (153), with case-control ratios of 1:22, 1:91, and 1:89 respectively.

A cohort comprising 160,000 white British samples was selected from the UKB WES dataset, and subsequently divided into three subgroups with sample size ratios of 1:1:1 and 4:3:2. Subsequently, Meta-SAIGE was employed to perform the analysis on these cohorts. The meta-analysis results were compared to the SAIGE-GENE+ results, which was performed on the whole 160,000 white British.

**Supplementary Figure 3** shows scatter plots that compare the results from Meta-SAIGE and SAIGE-GENE+ for type 2 diabetes, glaucoma, and ColCa. Meta-SAIGE successfully preserved the true signals as indicated by SAIGE-GENE+ and exhibited high correlations (**Supplementary Table 5**), without presenting any genes that were not significant in SAIGE-GENE+ results, but significant in Meta-SAIGE. It is noteworthy that the cohort-specific collapsing method also effectively maintained the signals from SAIGE-GENE+

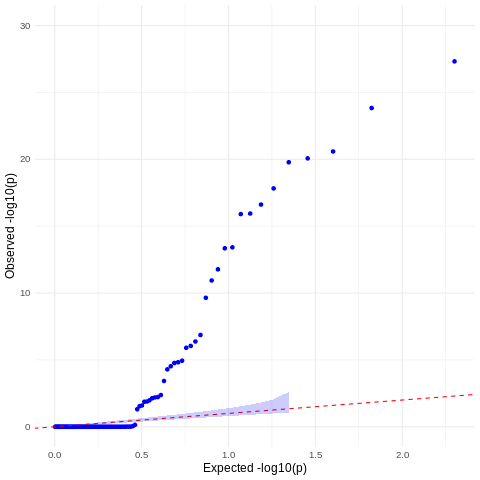

**Supplementary Figure 1.** QQplot for MetaSTARR results on a simulated phenotype with prevalence 1%. Repeated 100 times on FAM83F gene from 3 cohorts, which comprised 10,000 samples each.

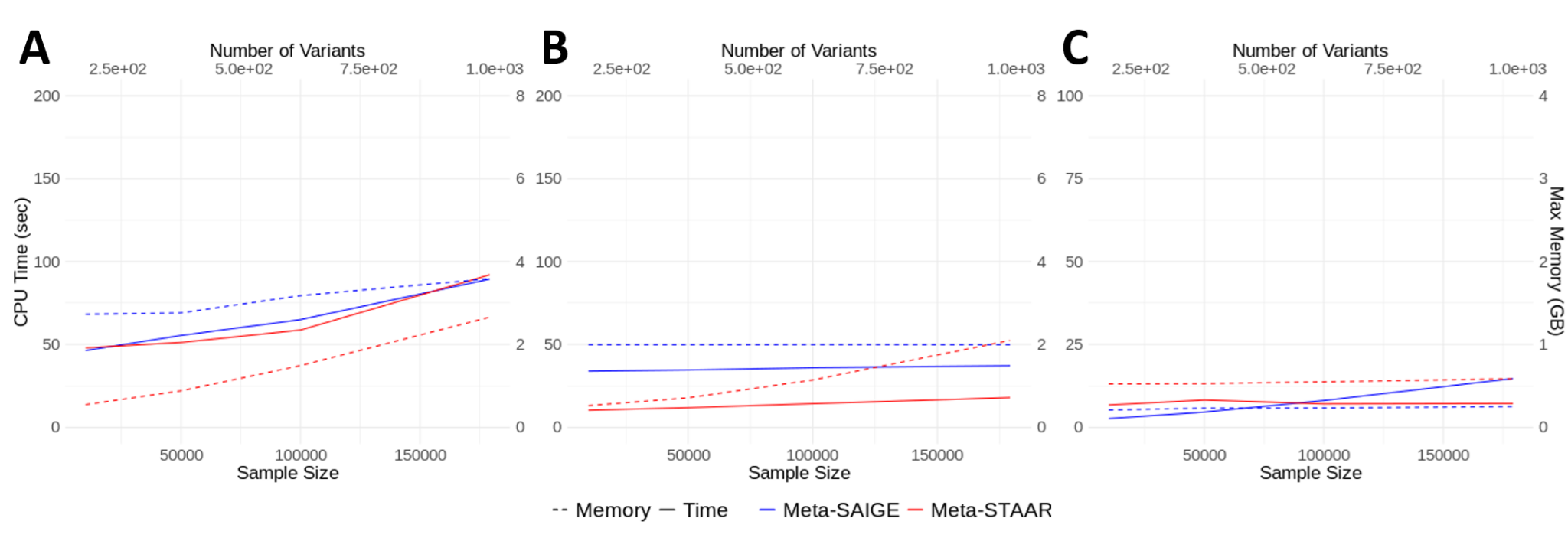

**Supplementary Figure 2**. Computational cost evaluation performed on a smaller gene, MSH6. A) Computational cost for input file generation. This step includes the null model fitting and summary statistics generation. B) Computational cost for LD matrix generation. C) Computational cost for meta-analysis step.

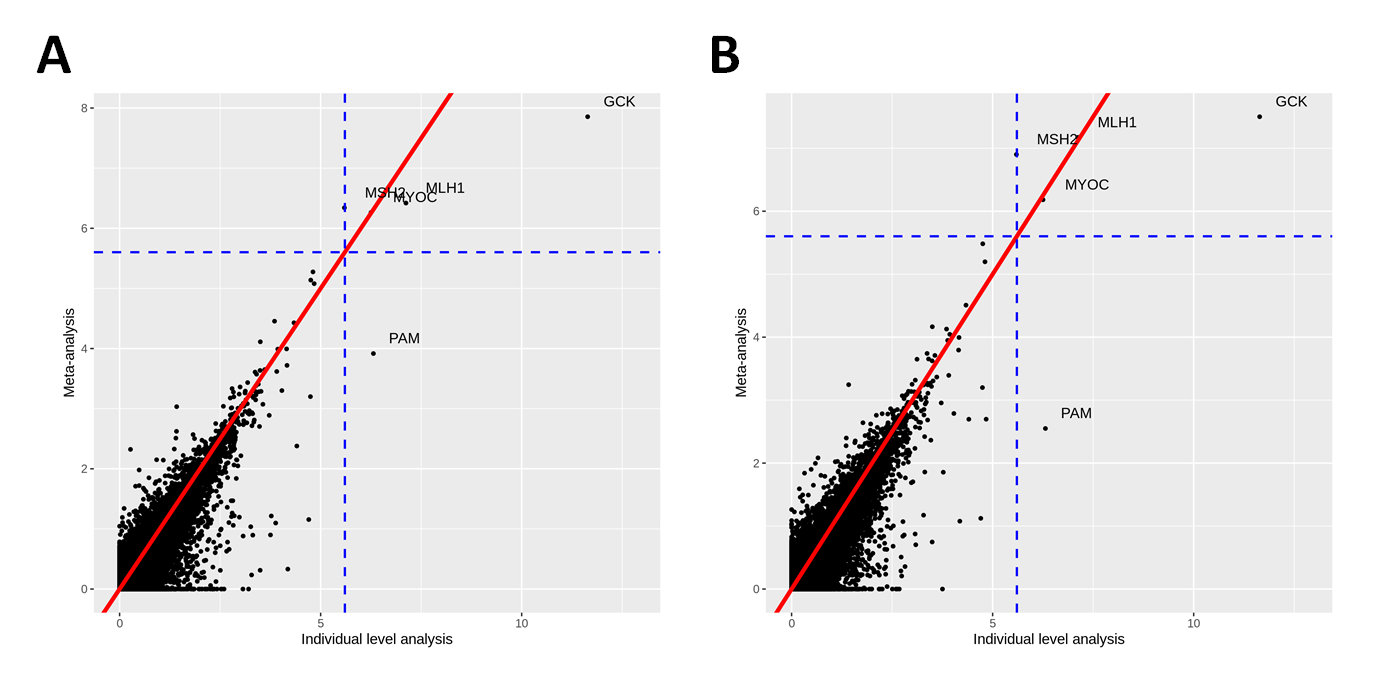

**Supplementary Figure 3.** Comparison of Meta-SAIGE to SAIGE-GENE+ using real data. The X-axis represents the -log10(p-value) of SAIGE-GENE+ and the Y-axis represents the -log10(p-value) of the meta-analysis. Significant genes are annotated in the plot. The blue lines represent the significance level ($2.5*10^{-6}$). A) Meta-SAIGE was performed on 1:1:1 sample size ratio. B) Meta-SAIGE was performed on 4:3:2 sample size ratio.

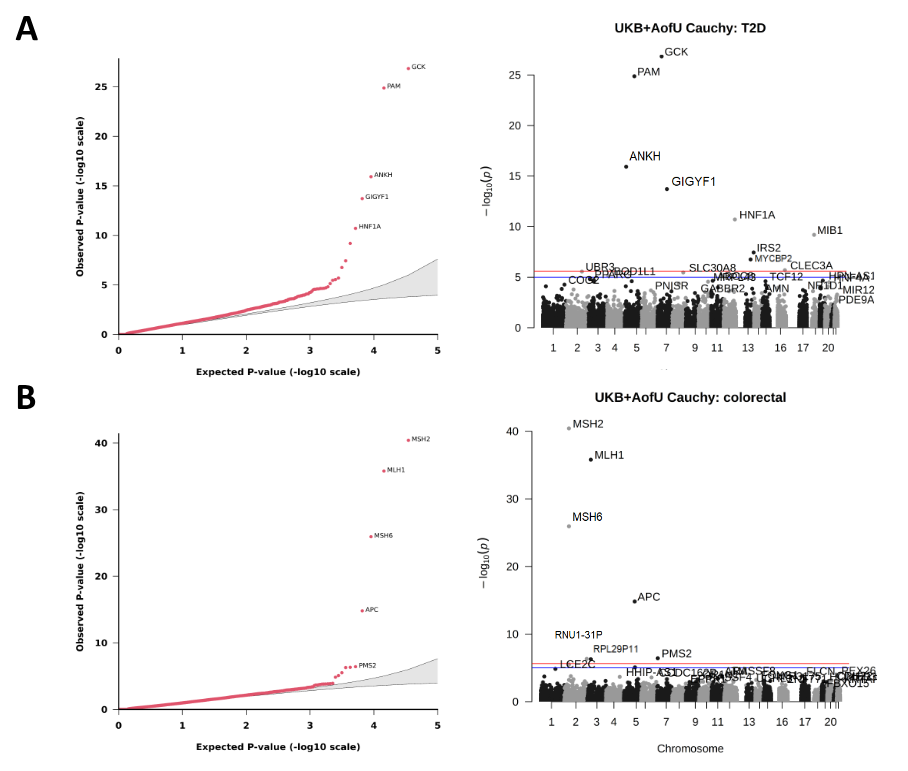

**Supplementary Figure 4**. Results for multi-ancestral analysis using AllofUs. A) Manhattan plot for type 2 diabetes (PheCode 250.2). B) Manhattan plot for colorectal cancer (PheCode 153)
